# The importance of time post-vaccination in determining the decrease in vaccine efficacy against SARS-CoV-2 variants of concern

**DOI:** 10.1101/2021.06.06.21258429

**Authors:** Yinon Bar On, Elad Noor, Noam Gottlieb, Alex Sigal, Ron Milo

## Abstract

With the development of high-efficacy vaccines against SARS-CoV-2, an urgent open question is whether currently available vaccines protect with similar efficacy against infection with SARS-CoV-2 variants of concern (VOC). Recent reports quantifying the extent by which VOC can evade vaccine immunity resulted in a range of estimates for the same VOC, which makes them difficult to interpret. One possible explanation for the discrepancies between different studies is an inconsistency in terms of the time post-vaccination of the sampled population. Here we present a model based on the observed correlation between antibody neutralization levels and vaccine efficacy, which demonstrates the impact of time post-vaccination on the comparison of the vaccine efficacy for VOC versus non-VOC infections. Our model predicts and exemplifies several possible consequences for vaccine efficacy in VOC infections: 1) a delay in the onset of vaccine efficacy against VOC; 2) a transient increase in susceptibility to breakthrough infection with VOC compared to non-VOC as a function of time after vaccination. We review preliminary data indicating that such phenomena are observed in studies of the B.1.1.7 and B.1.351 variants. We find that ignoring the strong dependence on the time post-vaccination can lead to contradictory reports of relative efficacy against VOC versus non-VOC, with implications on mitigation strategies against VOC and the design of vaccine efficacy studies.

## Introduction

The onset of global vaccination campaigns marks a significant milestone in the global effort to curb the spread of COVID-19. Current vaccines have different mechanisms for delivery of antigen (which with the exception of inactivated vaccines consists of SARS-CoV-2 spike), and with varying levels of efficacy, ranging from 60%-95% ^1^. A key determinant of the host immune response against viral infection is the production of neutralizing antibodies. Several recent studies have suggested a correlation between the level of neutralization elicited by different COVID-19 vaccines and their efficacy ^1,2^, at least in the early stages following vaccination.

Among the various SARS-CoV-2 variants that have emerged thus far, those associated with higher transmissibility, severity of disease, and escape from antibody responses to vaccines are of particular concern. These include variants first identified in the United Kingdom (B.1.1.7), South Africa (B.1.351), and Brazil (P.1). Specifically, VOC B.1.351 is capable of some immune evasion, with laboratory studies demonstrating a reduction in neutralization capacity of sera from vaccinated individuals as well as individuals naturally infected with a non-VOC virus ^3–9^. A dramatic decrease in neutralization capacity elicited by the AstraZeneca ChAdOx vaccine of the dominant B.1.351 variant in South Africa was associated with loss of vaccine efficacy during a phase 1b/2 clinical trial ^10^. The Johnson and Johnson adenovirus vaccine and the Novavax vaccine have also shown reduced efficacy in clinical trials conducted in South Africa during the time when the circulating variant was B.1.351 ^11,12^.

Vaccinated individuals tend to have less infections than unvaccinated individuals for all strains, but the relative protection may change between the VOC and the non-VOC, indicating partial immune evasion by the VOC ^13,14^. One approach to estimating vaccine efficacy against VOC is to compare efficacies between locales with different circulating variants. For example, in Israel, the Pfizer BNT162b2 vaccine roll-out was shown to be highly effective even though the B.1.1.7 variant was the dominant strain at the time ^15,16^. An alternative approach is to use case-control studies, where vaccinated individuals are matched with unvaccinated individuals with similar demographics and statistical tests are applied to examine the extent by which VOC infections are over-represented in the vaccinated population. A recent study using this methodology ^13^ indicated that B.1.351 is enriched in individuals vaccinated with the Pfizer BNT162b2 vaccine relative to non-VOC individuals, indicating a decrease in efficacy, although the results had wide confidence intervals due to the low number of cases. Another recent study suggested that the efficacy of the BNT162b2 vaccine against the B.1.351 variant 14 days after the second vaccine dose is about 75% (compared to the 95% reported for non-VOC strains) ^14^.

The increased interest in characterizing the extent of immune evasion by different VOC has resulted in several studies quantifying such evasion for different VOC and using different methodologies ^13,14,17^. However, these studies reach different quantitative conclusions that are challenging to reconcile. Most studies quantifying the extent of immune evasion by VOC do so at a specific time range post vaccination and, in many cases, attempt to characterize “fully immune” individuals, for which sufficient time has passed since their complete vaccination. However, studies define “full immunity” differently.

Here we investigate whether time post-vaccination is an important factor influencing the result of studies quantifying VOC immune-evasion. To demonstrate the utility of accounting for time post-vaccination, we present a toy model that quantifies the efficacy against VOC over time and shows that it is highly dependent on time post vaccination. We use this model to predict phenomena that can occur in studies of vaccine efficacy against VOC and discuss the implications of those predictions for the study of changes in vaccine efficacy against VOC.

## Results

Our toy model relies on two main components. First, we use a functional relationship between the neutralization level (denoted*n*) and vaccine efficacy (denoted*E*), as shown in Figure 1A. Similar to Khoury et al. ^1^, we define neutralization levels as the antibody neutralization level normalized to convalescent patients. This definition permits comparison across different measurement methodologies ^1^. Vaccine efficacy is defined as the reduction in the probability of infection in a vaccinated individual relative to an unvaccinated individual. We expect the relationship to be monotonic, with increasing neutralization levels associated with higher protection against infection. We use the relationship between neutralization levels and vaccine efficacy, as suggested in Khoury et al., as it is able to capture the variable effectiveness of different vaccines ^1^. We recognize that immune responses such as the T cell response likely play important roles in protection. We chose to focus here on neutralization as the correlate of protection because most of the effects of VOCs were measured in terms of the reduction in neutralization. The main results of our analysis can be directly generalized to the effects of VOCs on other correlates of protection such as the cellular response, as we explain in the discussion.

**Figure 1.**
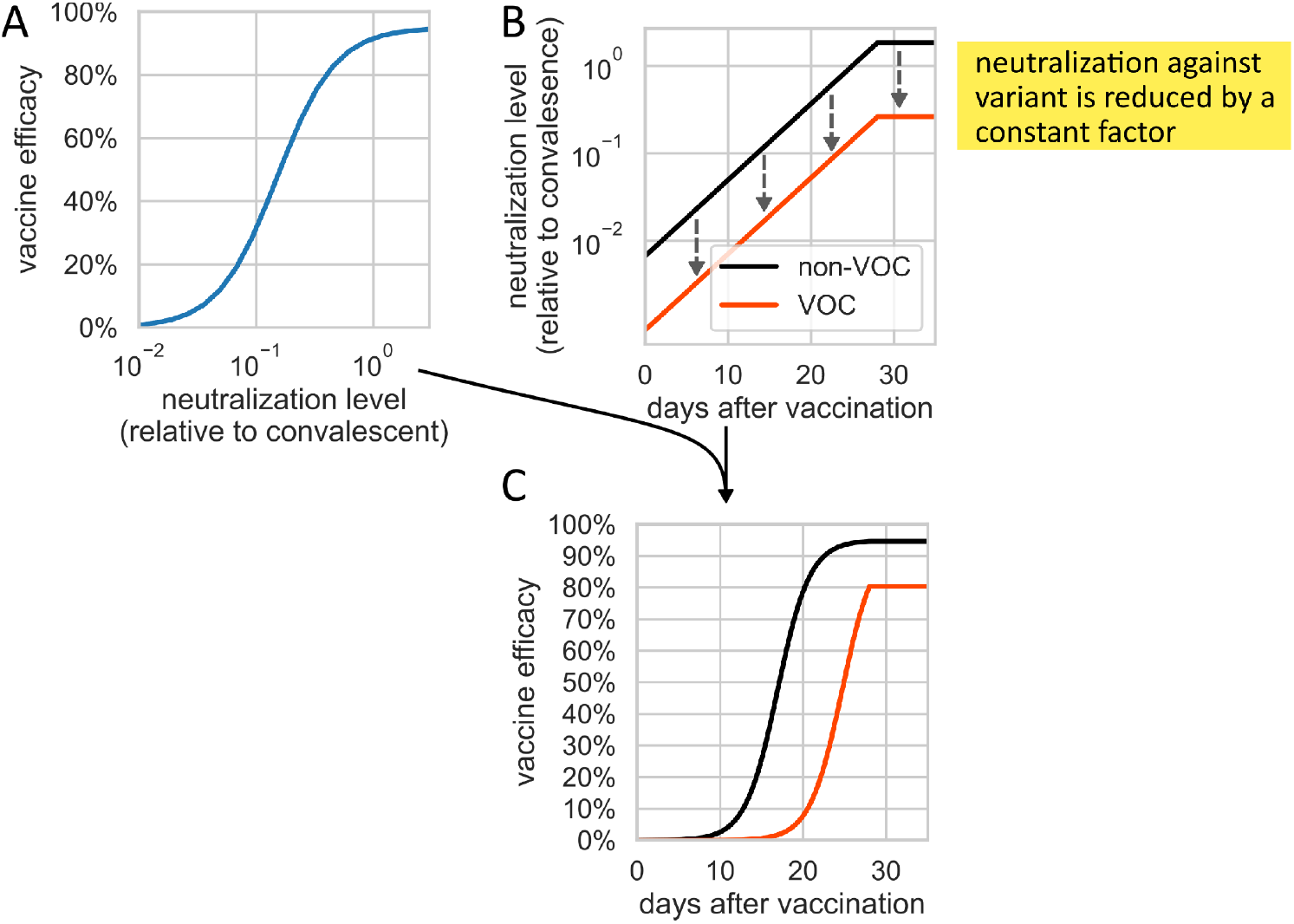
Schematic representation of the mathematical framework to model vaccine efficacy against VOC versus non-VOC. The model relies on the functional relationship between neutralization levels and vaccine efficacy (A) and the temporal profile of neutralization levels after vaccination (B). Using the functional relationship in A, we can convert neutralization levels into vaccine efficacy and create a temporal profile of vaccine efficacy as a function of time since vaccination (C). The parameter definitions and full details of the model are provided in the methods section and the SI. The parameter values used to generate the results are: N_fin_=4; d=7; K=1.7; E_max_=95%; n_50_=0.15.

We model the dynamics of the buildup of neutralizing antibodies in sera after vaccination. In the model, neutralization capacity starts below the detection limit and increases exponentially until it reaches a maximum persistent level about a week after the final vaccination dose ^18^. We use neutralization levels measured for the Pfizer BNT162b2 vaccine ^18^, yet similar observations hold for the Moderna vaccine and other single- and double-dose vaccines with appropriate changes in the parameter values. Over longer time periods (months to years), neutralization levels start to decrease ^1^. We assume initial neutralization levels (before vaccination) are negligible, such that *E* = 0, and after the first dose of the vaccine, neutralization levels rise exponentially until reaching the levels measured seven days after the second dose of the vaccine. Beyond that point, we assume neutralization levels stay constant in the short term. A schematic representation of the simplified temporal profile of neutralization levels is presented in Figure 1B.

Using the assumed functional relationship between neutralization levels and vaccine efficacy, we convert the temporal profile of neutralization levels into the corresponding temporal profile of vaccine efficacy (Figure 1C). We model vaccine efficacy against a VOC with immune evasion (such as B.1.351) using the same functional relationship, but assuming neutralization levels are reduced by a constant factor (e.g., 10-fold), based on observations in recent studies ^3–9^. The reduction in the neutralization level implies lower vaccine efficacy at each time point (Figure 1C). If a VOC strain does not evade the vaccine at all, but rather is concerning because of increased transmissibility, relative vaccine efficacy should be ≈1 for this variant.

This model makes several predictions. The first is that reduced neutralization levels could lead to a delay in the onset of vaccine efficacy against VOC. As can be seen schematically in Figure 1C, the reduction in neutralization levels against the VOC strain leads to decreased efficacy relative to the non-VOC strain at all time points (even when the efficacy after reaching maximal neutralization is similar as shown in Figure S1). Therefore, recently vaccinated individuals will reach a particular threshold level of vaccine efficacy at a later time post-vaccination than would be assumed based on data from non-VOC trials. Under parameter values within the ranges observed in the literature, which indicate a relatively small decrease in neutralization against the B.1.1.7 variant ^3–9^, the model produces a delay in vaccine efficacy, while keeping the final efficacy similar to that observed against non-VOC (Figure S1). Initial indications for such a phenomenon are found in the differences between the vaccine efficiency reported in clinical trials of the Pfizer BNT162b2 vaccine and the vaccine efficacy estimated in a roll-out of the vaccine, for example, in Israel ^15,16^. In clinical trials of the Pfizer BNT16b2 vaccine, significant protection against symptomatic infection reaches ≈90% as early as 14 days after the first Pfizer BNT162b2 vaccine dose. However, in later reports from Israel, despite a similar final efficacy to that of clinical trials, efficacy reached those levels only after the second vaccine dose ^15,16^. While the differences between the results of the clinical trials and the roll-out studies could stem from various sources, the results are consistent with the predicted delay in the onset of vaccine efficacy due to the high prevalence of the B.1.1.7 variant in Israel at the time. A similar pattern was also observed when studying efficacy against the B.1.1.7 variant in Qatar ^14^.

A second prediction offered by our model is that the relative susceptibility of the population to VOC (i.e. as compared to non-VOC strains) can vary non-monotonically over time. This can be seen as a transient peak in relative susceptibility to VOC in Figure 2. The relative susceptibility of vaccinated individuals to infection with VOC can be defined as the ratio between the fraction of the vaccinated population susceptible to a VOC strain, 1 − *E*_*V*_,and the fraction of the vaccinated population susceptible to non-VOC strains,1 − *E*. The transient peak appears as a result of the temporal progression of antibody buildup. Initially, both the non-VOC strain and the VOC strain have low neutralization levels, and thus low protection against infection (Figure 2A). Therefore, the susceptibility to infection with VOC and non-VOC is similar (Figure 2B). Shortly after the second vaccine dose, neutralization levels for the non-VOC strain increase to a degree that provides close-to-maximal protection, whereas the lower neutralization capacity against the VOC strain provides less protection (Figure 2). Therefore, there is an increase in the relative susceptibility of the vaccinated population to infection with VOC at these time points. In the following days, neutralization levels continue to increase for both the non-VOC and VOC strains, such that protection against the VOC further improves, decreasing the expected relative susceptibility of infected individuals to infection with the VOC compared with the non-VOC from its peak value (Figure 2B).

**Figure 2.**
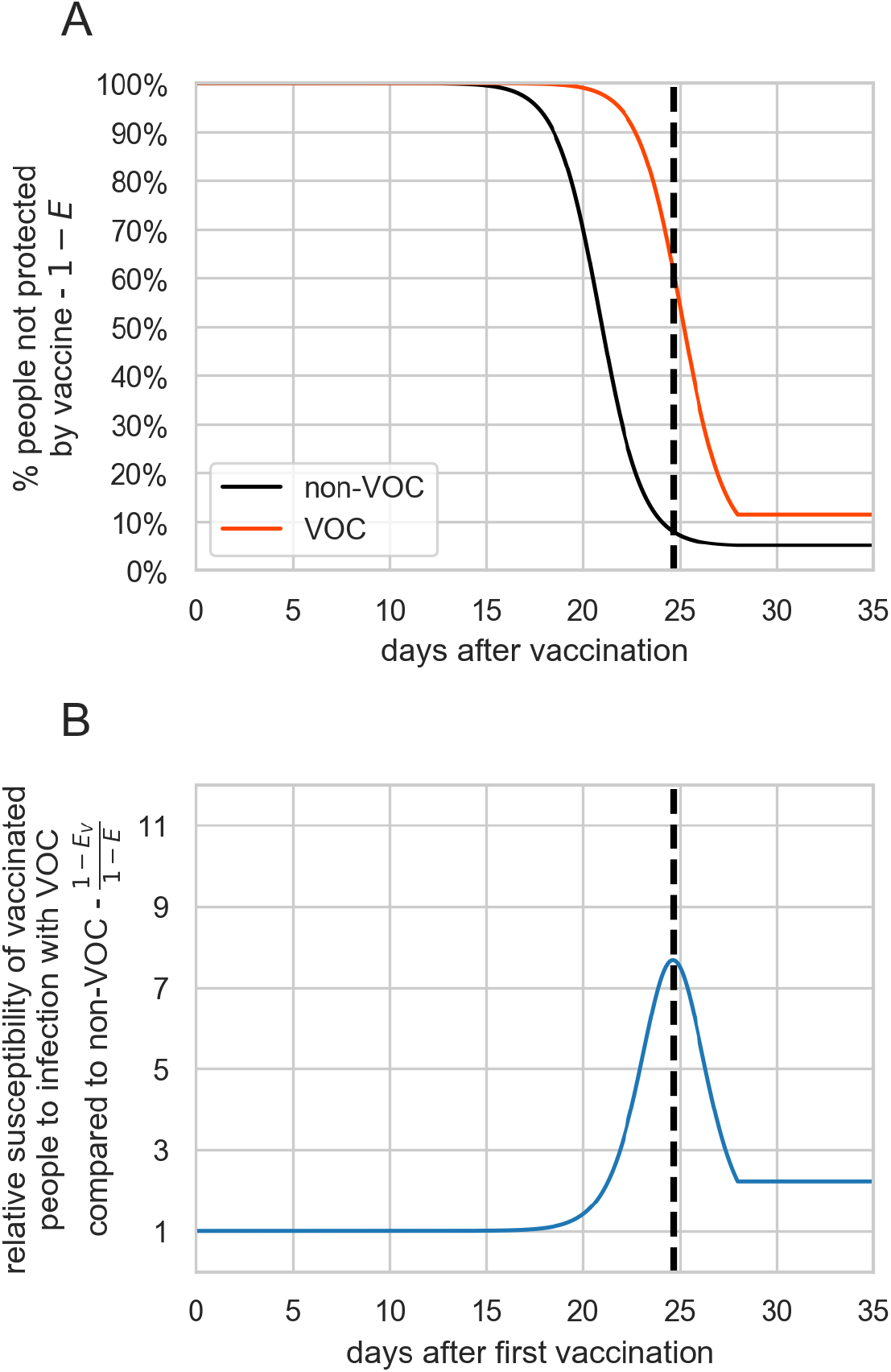
The relative susceptibility to infection with VOC compared with non-VOC in the vaccinated population. *A. The fraction of the vaccinated population not protected by the vaccine, for both the non-VOC strain (in black) and for the VOC strain (in orange) as a function of time since the first vaccine dose. B. The ratio between the susceptibility to infection with VOC and the susceptibility to infection with non-VOC in the vaccinated population, given by* 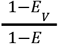 and plotted as a function of time since the day of vaccination. *The parameter values used to generate the results are: N_fin_=4; d=7; K=1.7; E_max_=95%; n_50_=0.15*

Initial evidence for the existence of such a transient peak can be found in a recent study that estimated the extent to which VOC B.1.351 is able to evade vaccine immunity ^13^. To estimate the extent of immune evasion by B.1.351, Kustin et al. studied pairs of vaccinated and unvaccinated infected individuals. Specifically, they investigated matched pairs in which the vaccinated and unvaccinated individuals were infected with different variants - termed discordant pairs. If the relative prevalence of VOC versus non-VOC among vaccinated people is higher than its relative prevalence among unvaccinated individuals, it provides evidence for an increase in susceptibility to infection with VOC compared with non-VOC in the vaccinated population. Overall, they observed that in eight out of the nine discordant pairs investigated, the vaccinated individuals were infected with the B.1.351 strain, and only in one case was the unvaccinated individual infected with the B.1.351 strain, suggesting a possible increase in the relative susceptibility to infection with VOC compared with non-VOC in the vaccinated population. However, even though Kustin et al. sampled an equal number of pairs in the first and second weeks after the second vaccine dose, all 8 discordant pairs in which the vaccinated individuals were infected with the B.1.351 strain were infected in the first week after the second dose, and none were infected in the second week. This suggests the existence of a transient peak in relative susceptibility in the first week after the second dose of the vaccine, as predicted by the model.

## Discussion

We present a toy mathematical model that builds upon a previously-described functional relationship between vaccine efficacy and neutralization levels ^19^. To this model, we add the temporal dynamics of neutralization levels observed in immunized individuals, as well as data regarding the decrease in neutralization levels for VOC strains of the virus. By combining these data sources, we are able to model vaccine efficacy against VOC. The model predicts that the relative efficacy of VOC compared with non-VOC is not constant over time and could potentially be non-monotonic, with a transient decrease in relative efficacy and a peak in relative susceptibility.

Beyond these predictions, our analysis has possible implications for vaccine efficacy against VOC in the long term. The long-term efficacy of currently-available vaccines is not well characterized. If it eventually wanes, neutralization levels will eventually drop back to the levels observed in the days following the second dose. If the temporal profile of relative susceptibility of vaccinated people to infection with VOC compared with non-VOC exhibits a transient peak (Figure 2B), then the decrease in neutralization levels could lead to increased susceptibility to infection by VOC in the long term.

We note that our model does not aim to provide a mechanistic depiction of the way in which VOC evade the immune system. We modelled the dynamics of antibody neutralization and vaccine efficacy in the simplest manner possible to help build intuition of how time post-vaccination plays a role in determining the efficacy against VOC.

A key simplification our model makes is that we chose to focus on antibody neutralization as the main correlate to protection against infection. While there is a strong correlation between final neutralization levels after vaccination and vaccine efficacy ^2,19^, there are other mechanisms of protection that are not based on antibody neutralization ^20^. One can generalize the model to consider the overall immune response to infection rather than neutralization levels specifically. As long as the general temporal dynamics of those responses is similar, and VOC cause a decrease in the effectiveness of the immune response, the qualitative results of the model would remain unchanged. An assumption made in constructing the model is that the functional relationship observed by Khoury et al. between maximal neutralization levels for each vaccine and vaccine efficacy also holds during the buildup of neutralizing antibodies in sera after vaccination. We also assume that the same functional relationship described for non-VOC also holds for VOC, only with a reduced neutralization level against VOC.

In terms of finding supporting evidence for the predictions, there are currently significant uncertainties in estimating the efficacy of vaccines against VOC as a function of time from empirical data. For example, the numbers supporting the immune evasion of the B.1.351 variant are very small, with the results of the study containing a large degree of uncertainty^13^. Moreover, almost all the information about breakthrough cases infected with the B.1.351 variant are confined to the first week after the second dose of the vaccine, leaving even greater uncertainty as to the vaccine efficacy against B.1.351 in later weeks.

The results presented here may indicate that time post-vaccination needs to be considered in the measurement of vaccine efficacy, and results need to be collected and stratified by timing post vaccination. Disregarding the strong dependence on the time post-vaccination can lead to contradictory conclusions on the relative efficacy against VOC versus non-VOC as evident in one study showing a large effect and another showing a small effect ^13,14^. While specific quantitative predictions of the presented model depend on the specific parameter values chosen, the general finding that the dynamics of the immune response could have significant implications for the observed relative vaccine efficacy against VOC holds true under a wide range of conditions (Figure S4). Real-world measurements are needed to fully validate these predictions. Measuring the time-dependence of relative vaccine efficacy against VOC and, more specifically, the degree of immunity in vaccinated individuals is an important effort that will require more exact control of time post-vaccination in future studies. A better understanding of how the prevalence of emerging VOC in the vaccinated infected population changes as a function of time since vaccination could help us understand both the short- and long-term implications of emerging VOC on the spread of COVID-19 and determine the need for modified vaccines to address them.

## Methods

### The relationship between antibody neutralization levels and vaccine efficacy

Our mathematical framework relies on the foundation laid by Khoury et al._-_^1^, who assume a quantitative relationship between neutralization levels and protection against infection at the individual level. To allow for generalization of the framework across the different types of neutralization assays used, the model normalizes the neutralization levels of vaccinated people to those of convalescent patients measured using the same method/in the same study. The positive correlation observed between neutralization levels and vaccine efficacy monotonically increases and is sigmoidal (S shaped) ^2,19^. This basic functional relationship between neutralization level and protection at the individual level can be represented as a hyperbolic transformation of the neutralization levels (to the power of k), which implies the following relationship:

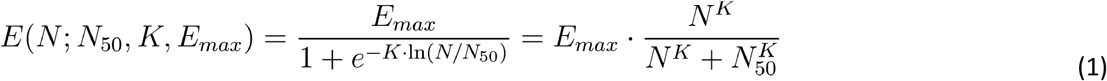

where *E* is the vaccine efficacy against infection, *E*_*max*_ is the maximal efficacy, *N* is the normalized neutralization titer, *N*_50_ is the normalized neutralization titer at which protection reaches one half of the maximal efficacy, and *K* is the steepness of the curve. Both *N*and *N*_50_ are given in units normalized to the neutralization level observed in convalescent patients, though we note that only their ratio, which is unitless, affects the results. Note that, unlike the notation in Khoury et al._-_^1^ where *n* represented the log_10_-transformed normalized neutralization titers, we use the untransformed titers directly (*N)*, so that *N* = 10^*n*^. As a consequence, the steepness *K* is scaled relative to the *k* in Khoury et al._-_^1^, i.e. *K* = *k*/*ln*(10).

We model how a new variant evades an immune response by a decrease in neutralization levels against it. We denote the degree of decrease in neutralization levels as *d* > 1. Thus, the vaccine efficacy against the infection by the new variant at the level of an individual person is given by:

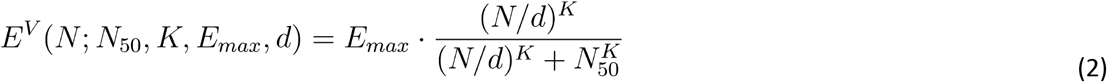

where *E*^*V*^ is the vaccine efficacy against the new variant. The difference between neutralization levels and vaccine efficacy for the VOC and non-VOC is described schematically in Figure 1. For the shown example, we use *d* = 10 and *E*_*max*_ = 95%.

### Parameter values used in the analysis

We applied the mathematical framework described above to study the temporal profile of relative vaccine efficacy against two VOC - the B.1.1.7 strain and the B.1.351 strain. In both cases, the final neutralization level we used was 4-fold higher than the neutralization level of convalescent patients. This level is within the range reported in the literature ^18,21^, i.e., about 2-4 fold higher than the neutralization levels measured for convalescent patients in the same study (depending on patient age). For the value for *N*_50_,we used 0.15, which is within the range estimated by Khoury et al. ^1^ (i.e., about one seventh of the neutralization level observed in convalescent patients is correlated with 50% efficacy). In our analysis, we used 1.7 as the value for*K* which is within the range reported in Khoury et al. (95% range of 1.0-1.9) ^1^, We chose the maximal vaccine efficacy *E*_*max*_ to be 95%. We used the decrease in neutralization levels *d* to be 2 for B.1.1.7, and 7 for B.1.351. Both values are within the range reported in recent neutralization assays ^3–9^. We emphasize that values measured *in vitro* (*d*) might change *in vivo* and the model used is an extreme simplification of the biological system, and we thus consider the parameters values used as a way to exemplify the possible phenomenon rather than a representative summary of the literature values. As a result we choose some parameter values to deviate from the most commonly cited values, yet still within reported ranges.

## Data Availability

All data used in the paper is publicly available in the literature

## Acknowledgments

We thank Avi Flamholz, Laurence Freedman, Jonathan Gershoni, Lior Greenspoon, Itay Hazan, Yoram Hemo, Amit Huppert, Micha Mandel, Natalie Page, Shmuel Ohayon, Ron Sender, Mike Springer and Adi Stern for valuable insights throughout this study.

## Supplementary Information

### Incorporation of variability in neutralization levels across the population

The simple model used in the main text relies only on the mean neutralization level across vaccinated individuals. In reality, neutralization levels vary between individuals. To take this variability into account, we follow Khoury et al. ^1^ and assume that neutralization levels are log-normally distributed across the population, meaning that the log-transformed neutralization levels are normally distributed. To incorporate this variability, we use equation 1 to model vaccine efficacy at the individual level, and calculate the population-level efficacy as the expectancy of the efficacy across the population:

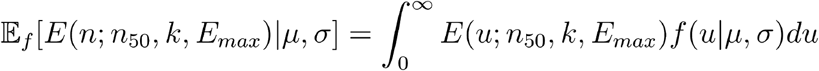

where *E* is the population-level efficacy against infection, *f(u*|*μ,σ)* is the probability density function of a log-normal distribution with *μ* and *σ* being the mean and standard deviation (respectively) of the log-transformed neutralization levels across the population.

The efficacy against the variant strain at the population level is similar to that for the non-VOC strain, but is derived using equation 2 instead of equation 1 for the individual-level vaccine efficacy:

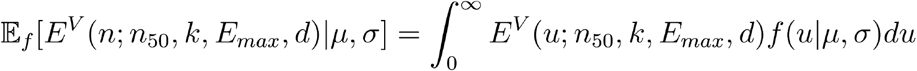

Using this more general framework, we perform the same type of analysis and arrive at similar results to those shown in the main text (presented in Figure S2-S3), taking a variance in neutralization levels within the range reported by Khoury et al. ^1^ (*σ*=0.3).

**Figure S1.**
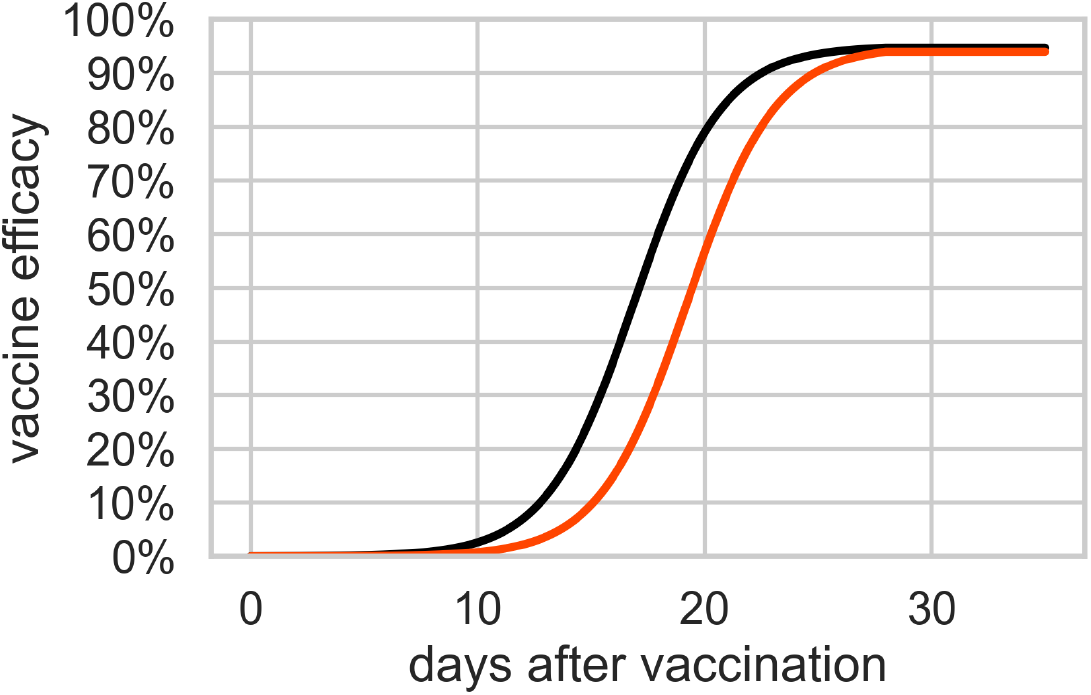
Estimates of the vaccine efficacy against non-VOC and the B.1.1.7 variant. We use the same framework and in Figure 1 but with different parameter values for d. The parameters used for this analysis are: N_fin_=4; d=2; K=1.7; E_max_=95%; n_50_=0.15.

**Figure S2.**
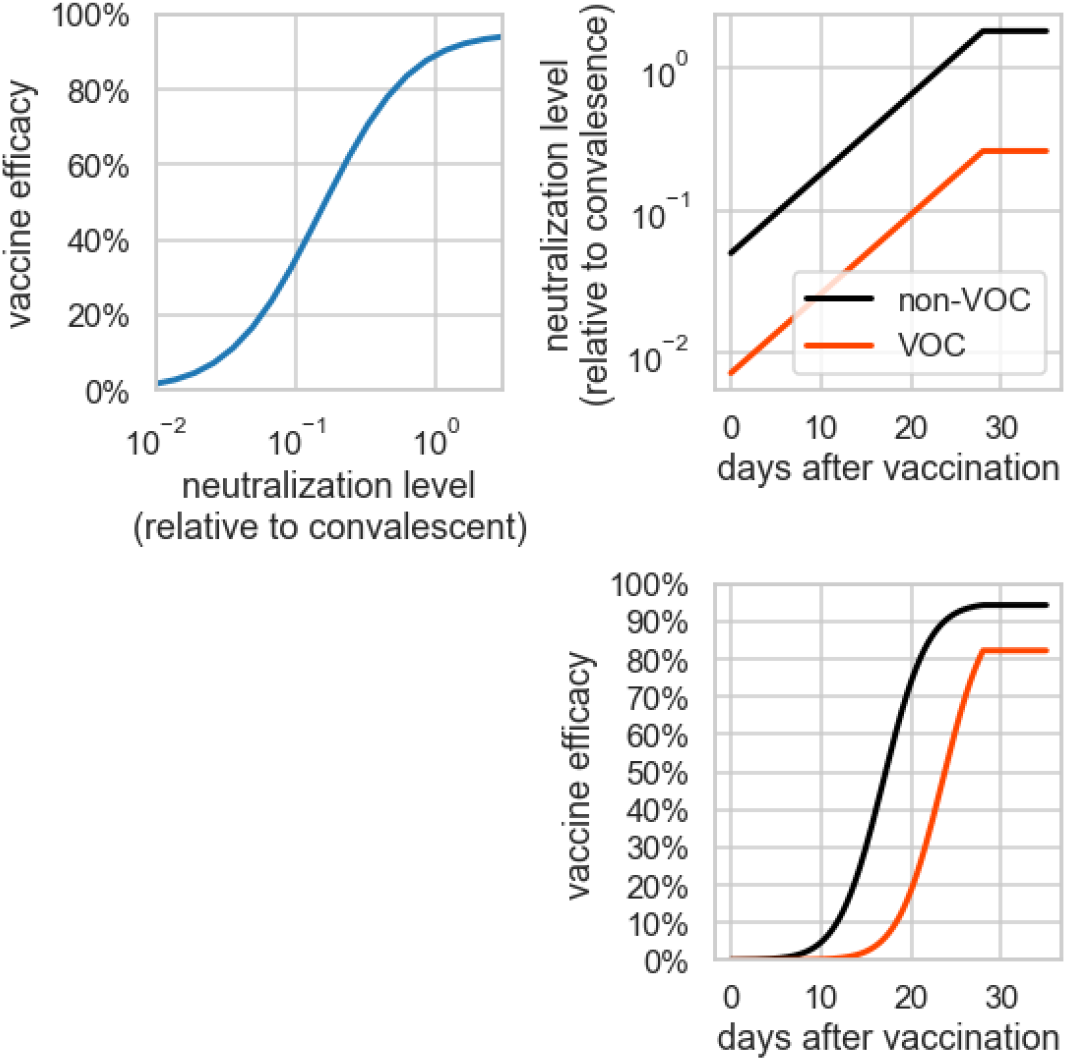
The effect of adding variability in neutralization levels on the results of Figure 1. The figure is the same as Figure 1 in the main text, except that a standard deviation of 0.3 log_10_ transformed neutralization levels is used. The other parameter values used are N_fin_=4; d=7; K=1.7; E_max_=95%; n_50_=0.15.

**Figure S3.**
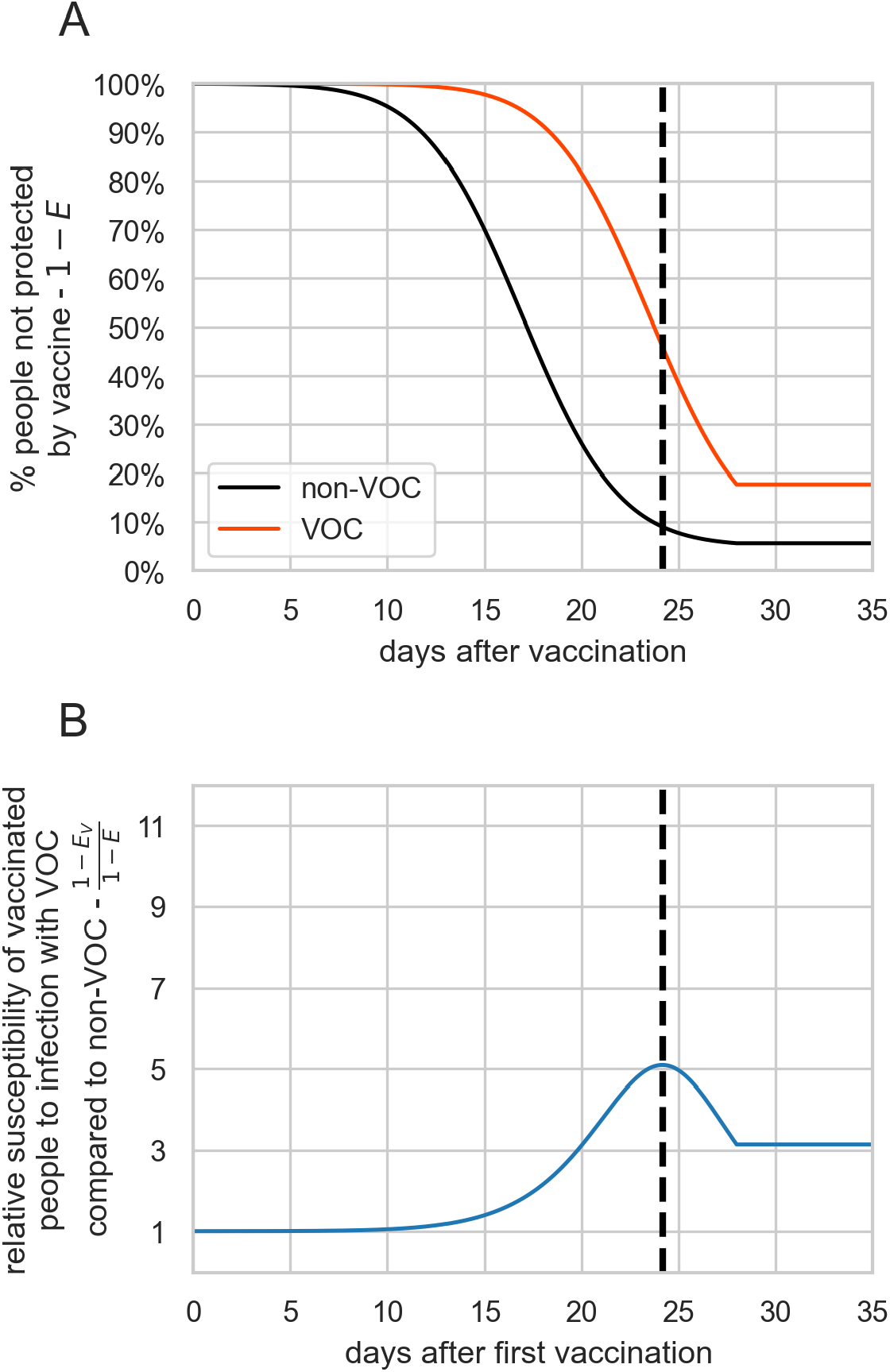
The effect of adding variability in neutralization levels on the results of Figure 2. The figure is the same as Figure 2 in the main text, except that a standard deviation of 0.3 log_10_ transformed neutralization levels is used. The other parameter values used are N_fin_=4;d=7;K=1.7;E_max_=95%; n50=0.15.

**Figure S4.**
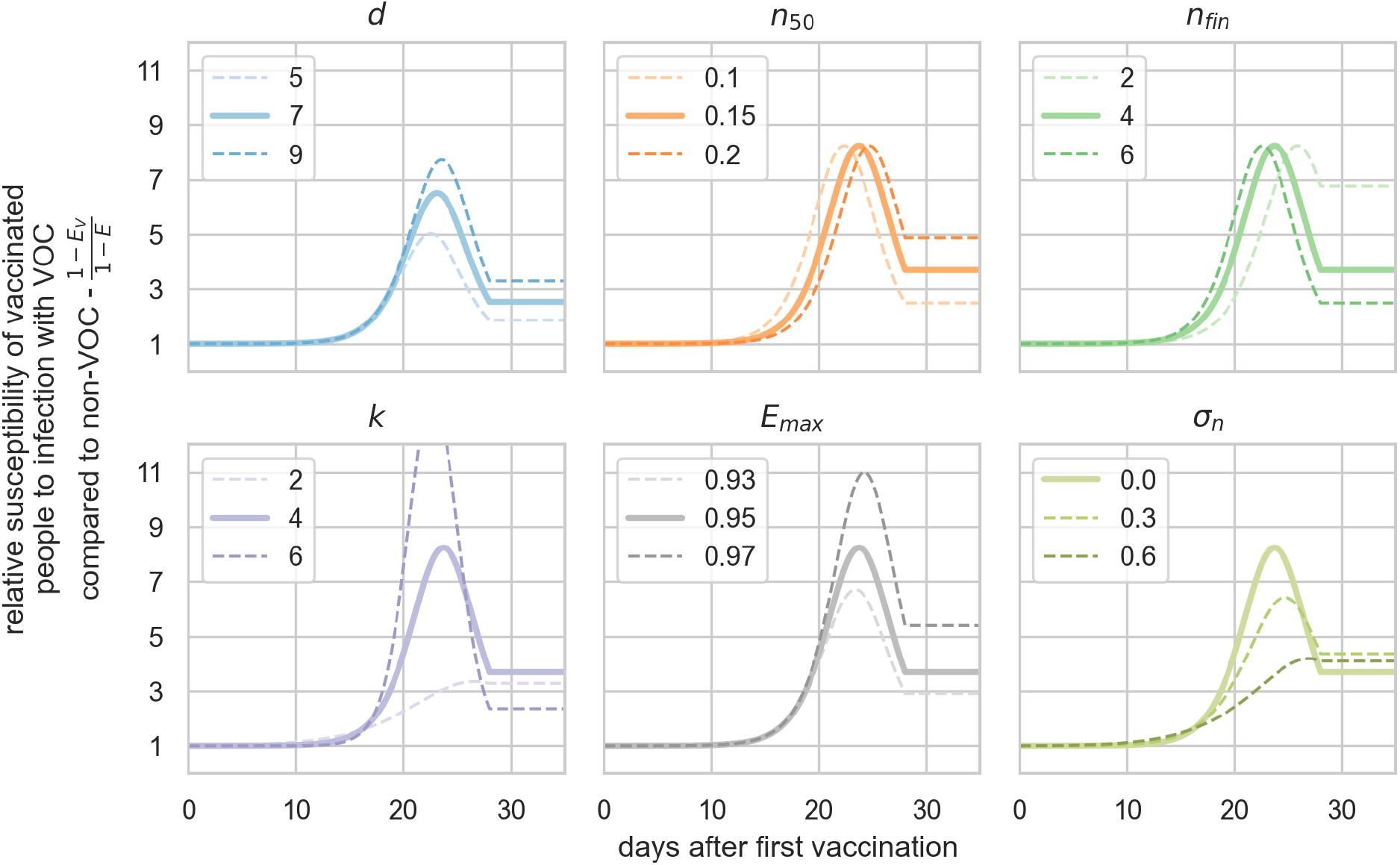
Sensitivity analysis for the time dependence of the relative susceptibility of vaccinated people to infection with VOC compared to non-VOC. In each panel, we change the value of one of the model parameters to test its influence on our results. For each parameter, the middle value represents the value used in our main analysis, except in the case of σ_n_ (where the default value is 0).

